# How it begins: Initial response to opioids strongly predicts self-reported opioid use disorder

**DOI:** 10.1101/2025.03.21.25324409

**Authors:** Jean Gonzalez, Vinh Tran, John Meredith, Ivonne Xu, Ritviksiddha Penchala, Laura Vilar-Ribó, Natasia Courchesne-Krak, Daniel Zoleikhaeian, Matthew McIntyre, Pierre Fontanillas, Katelyn K. Bond, Eric O. Johnson, Alvin D. Jeffery, James MacKillop, Carla Marienfeld, Harriet de Wit, Abraham A. Palmer, Sandra Sanchez-Roige

**Affiliations:** Department of Psychiatry, University of California San Diego, La Jolla, CA, USA; 23andMe, Inc., Sunnyvale, CA, USA; GenOmics and Translational Research Center, RTI International, Research Triangle Park, NC, USA; Fellow Program, RTI International, Research Triangle Park, NC, United States; Vanderbilt University School of Nursing, Nashville, Tennessee, USA; Department of Biomedical Informatics, Vanderbilt University Medical Center, Nashville, Tennessee, USA; Peter Boris Centre for Addictions Research, St. Joseph’s Healthcare Hamilton, Hamilton, ON, Canada; Department of Psychiatry and Behavioral Neurosciences, McMasters University, Hamilton, ON, Canada; Department of Psychiatry and Behavioral Neuroscience, University of Chicago, Chicago IL, USA; Institute for Genomic Medicine, University of California San Diego, La Jolla, CA, USA; Division of Genetic Medicine, Vanderbilt University Medical Center, Nashville, TN, USA

**Author notes:** **Co-corresponding authors:** Dr. Abraham A. Palmer, Dr. Sandra Sanchez- Roige.

## Abstract

**Background:** Opioid use disorder (**OUD**) is a major public health crisis. Patients’ initial exposure to opioids often comes from prescribed medications. Predicting which of these patients will develop OUD remains challenging. Prior evidence from various substances suggest that initial subjective responses influence addiction risk, however these studies have used relatively small cohorts and have not led to the development of widespread tools to predict OUD risk.

**Methods:** We used a cohort of 141,897 adult research participants to perform a retrospective observational study of self-reported subjective responses to prescription opioids. We collected demographics, subjective positive (e.g., euphoria), subjective negative (e.g., nausea), and analgesic responses as well as self-reported OUD.

**Results:** Positive subjective effects, particularly “Like Overall”, “Euphoric”, and “Energized”, were the strongest predictors of OUD. For example, the odds-ratio for individuals responding “Extremely” for “Like Overall” was 36.5. The sensitivity and specificity of this single question was excellent (ROC=0.87). Negative effects and analgesic effects were much less predictive. We developed a two-question decision tree (“*When you first took opioid pain medication, to what extent did you like the way they made you feel overall?*” and “*When you first took opioid pain medication, to what extent did you experience an unpleasant itchy feeling?”*), that can identify a small high-risk subset with 78.5% prevalence of OUD and a much larger low-risk subset with 1.2% prevalence of OUD.

**Conclusions:** Screening for subjective responses can identify high-risk individuals who would benefit from tailored interventions.

## INTRODUCTION

The opioid epidemic remains a major public health crisis, with more than 100,000 overdose deaths per year in the United States alone. Opioids are often first encountered in medical settings for pain management. A substantial minority of individuals go on to develop opioid use disorder (**OUD**).^1^ Early identification of individuals with heightened risk could guide clinical decision-making and prevention strategies, yet current tools for risk stratification are limited in both scalability and predictive performance.^2,3^ Thus, there is an urgent need to identify which of the patients being considered for an opioid prescription are at greatest risk for OUD.

A growing body of evidence suggests that an individual’s initial subjective response to a drug may shape subsequent use trajectories and risk for addiction.^4–11^ In alcohol research, individuals who are more sensitive to alcohol’s negative effects are at decreased risk for future alcohol use disorder.^10,12^ In addition, individuals who are more sensitive to the positive effects of alcohol, stimulants, and cannabis are at increased risk for future substance use disorder.^8,13^ There is also evidence that initial positive subjective responses may be a risk factor for OUD.^14– 18^ However, existing studies are small, laboratory-based, and have been ascertained in ways that limit their generalizability.

To address these gaps, we analyzed data from 141,897 23andMe research participants who completed a retrospective, online survey about their first experience with prescription opioids.^19^ We asked participants to recall their initial subjective responses to prescription opioids. We also asked participants to self-report lifetime OUD status, and cross-referenced their responses with 9 questions pertaining to problematic opioid use to reduce potential sample misclassification. Using a combination of machine learning classifiers and a decision tree framework, we developed a concise screener that stratifies risk for OUD.

## METHODS

### Study Design and Cohort Acquisition

We used a cohort of 198,906 research participants that were drawn from the customer base of 23andMe, Inc., a consumer genetics and research company. Participants provided informed consent and volunteered to participate in the research online, under a protocol approved by the external AAHRPP-accredited Salus IRB (https://www.versiticlinicaltrials.org/salusirb). This survey was active from May 23, 2023 to May 15, 2025, it was advertised to all 23andMe research participants through email and available through their online portal. No compensation was provided.

Only participants who endorsed the statement, *“Have you ever taken any opioid pain medications or narcotics regardless of whether they were prescribed to you or not? For example, codeine, hydrocodone, oxycodone, or morphine”*, were included in the study. These participants were asked 11 questions about their initial subjective responses to opioids, 9 questions about problematic opioid use, and 1 question about self-reported diagnosis of OUD (**Table S1**).^19^

The 11 questions captured initial subjective effects including 1 question about analgesia (e.g. “*When you first took pain medication, to what extent did you feel less pain*?”), 5 questions about positive effects (i.e. “Euphoric”, “Energized”, “Normal”, “Relaxed or Calm”, “Like the way you feel overall” [“Like Overall”]) and 5 questions about negative effects (i.e., “Nauseated”, “Dizzy”, “Tired”, “Constipated”, “Itchy”). Response options included: “Not at all”, “Mildly”, “Moderately”, “Very much”, “Extremely”. These questions were adapted from the Opioid Checklist^13^, the Addiction Research Center Inventory^14^, and the History of Opioid Medical Exposure^15^.

9 additional questions were intended to capture key aspects of lifetime problematic opioid use, which were summed to produce a score that ranged from 0 to 27 (see **Table S1** for the full set of questions). These questions were adapted from the Non-Medical Opioid Use scale, Opioid Risk Tool^20^ (**ORT**) Current Opioid Misuse Measure^21^, Prescription Opioid Misuse and Abuse Questionnaire^22^, Prescription Opioid Misuse Index^23^, and Addiction Severity Index^24^.

We included 1 question that asked *“Have you ever felt that you might have a problem with, or have you ever been diagnosed with or treated for, addiction to opioids?”*, with response options including *“Yes, I felt I might have a problem but have never been diagnosed or treated”, “Yes, I have been diagnosed or treated”*, or *“No”*.^25^ We defined OUD to include both self-diagnosis and self-reported clinical diagnosis because OUD is known to be underdiagnosed.^26,27^ We performed our key analysis for only the self-diagnosed or only the self-reported clinical diagnosis and confirmed that they behaved similarly (**Figure S1**).

To minimize misclassification errors, participants were also excluded if their problematic opioid use score was discordant with their response to the self-reported lifetime OUD question (**Figures S3, Figure S4**). Specifically, we excluded participants who indicated they had OUD but whose problematic opioid use score was below 3 (*n* = 2,442, 1.2%). We also excluded participants who responded that they did not have OUD but whose problematic opioid score was above 5 (*n* = 9,243, 4.6%). These values were chosen empirically based on the distribution of the problematic opioid use scores (see **Supplemental Methods** for details).

Finally, demographic information (**Table 1**) that was available from 23andMe included age, biological sex, education, income, and genetically defined ancestry.^28^ We also asked participants about the age at first opioid use. From this, we calculated the average time between first use and survey completion, which was 24.4 years (SD = 13.3).

In addition to excluding subjects due to the discordance between self-reported OUD and problematic opioid use score (see above), participants were also excluded if they had any missing responses (*n* = 36,568, 18.4%), for inattention, age at first use under 12 years old (*n* = 7,082, 3.6%), current age above 90 years old (*n* = 306, 0.2%), and if age at first use was older that current age (*n* = 39, 0.1%). The number of participants excluded at each step is described in **Supplemental Methods**; after all exclusions, 141,897 participants remained (**Figure S2**).

### Statistical Analyses

We used responses to subjective effects questions to predict OUD status with three classifiers: Logistic Regression, XGBoost, and Random Forest and aggregated the results to form an ensemble model.^29,30^

We evaluated model performance via area under the receiver operating characteristic curve (**AUC-ROC**), summarizing the model’s ability to discriminate between OUD and non-OUD; precision, measuring the proportion of correct positive predictions; recall (true positive rate), quantifying the proportion of actual positives correctly identified; and the F1 score, the harmonic mean of precision and recall. We also plotted precision-recall (**PR**) curves to visualize the trade-off between precision and recall, with the area under the PR curve (**PR-AUC**) providing an overall summary. To ensure robustness, we used 5-fold cross validation (see **Supplemental Methods**).

We assessed feature importance using SHapley Additive exPlanations^31^ (**SHAP**) values, which were normalized and averaged across classifiers. Odds ratios (**ORs**) were calculated for a subset of questions, comparing participants who selected “Extremely” to those who selected any of the other options (“Very much”, “Moderately”, “Mildly”, or “Not at all”).

We trained a decision tree classifier using *sklearn*^30^ to identify the minimal set of questions relevant to OUD that could be used to stratify risk for OUD. We used 5-fold cross validation and found that the same questions were selected in all 5 instances (see **Supplemental Methods**).

## RESULTS

The sample was 70.42% female with a mean age of 52.99±0.04 years. 10.11% of the sample had a household income of $100,000 or more. 87.41% of the sample had at least some college education. Demographic and other information about the cohort is presented in **Table 1** and **Table S2**.

**Figure 1a** compares the mean responses and distributions of subjective effects at first opioid use for individuals self-reporting OUD versus those who did not self-report OUD. Questions are sorted by the degree to which individuals with OUD scored higher than individuals without OUD. Positive subjective effects, such as “Like Overall”, “Euphoric”, and “Energized”, were much higher in individuals with OUD, suggesting that sensitivity to the hedonic effects of opioids is a risk factor for developing OUD. In contrast, for the negative subjective effects, the differences between the groups were lower in magnitude. Furthermore, the direction of the negative effects was not consistent (**Figure 1b**). More specifically, “Itchy”, “Constipation”, and “Nauseated” were higher in the individuals with OUD, whereas “Tired” and “Dizzy” were lower in the individuals with OUD, suggesting that the negative subjective effects of opioids are a less reliable risk factor for OUD. Finally, “Less Pain” was only slightly higher in individuals with OUD, suggesting that the analgesic effects of opioids, which could be construed as positive, are not a strong risk factor for OUD. Full descriptive statistics are provided in **Table S3**. Correlation estimates for subjective effects are shown in **Figure S5**.

**Figure 1.**
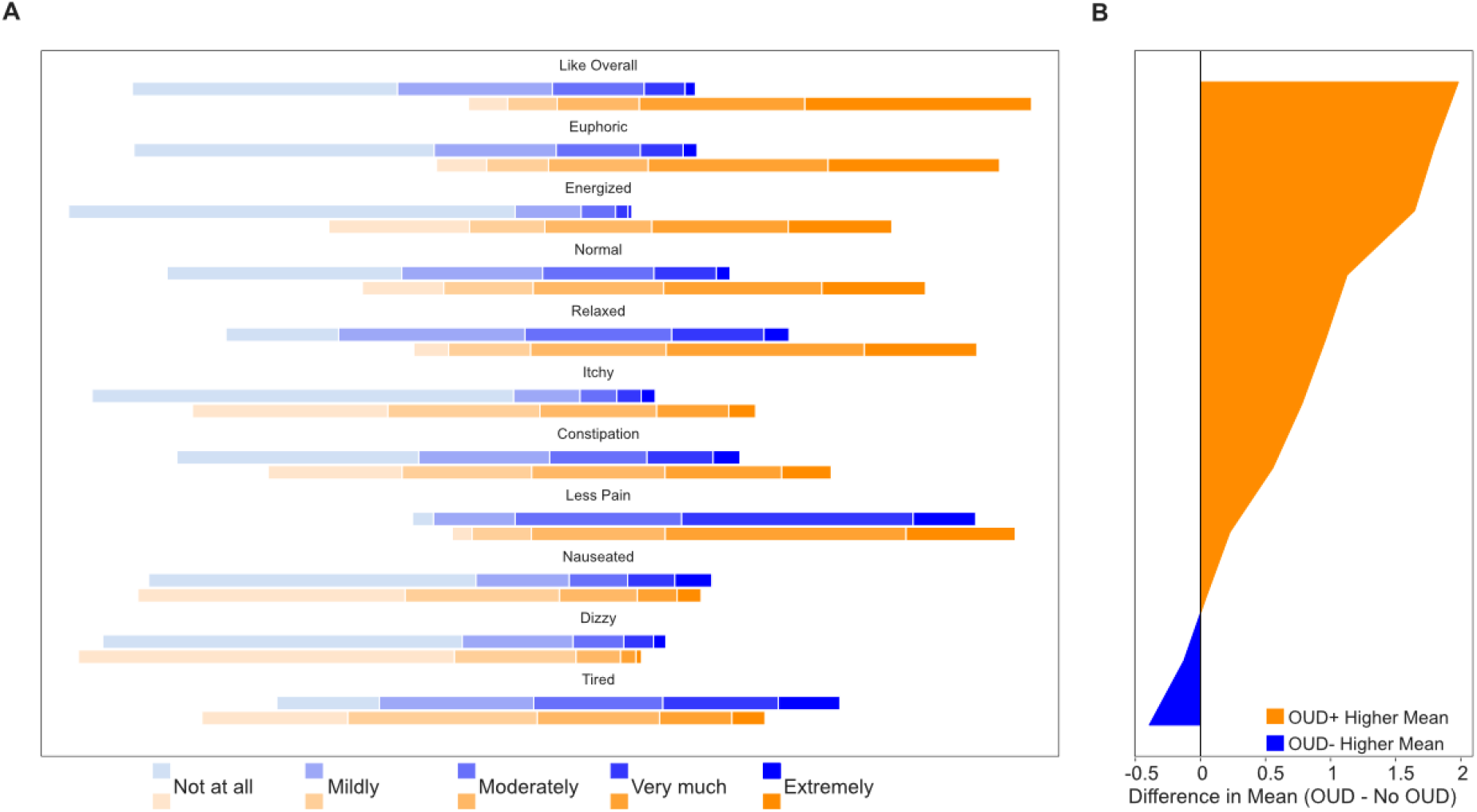
Distribution of subjective effects reported at first use for individuals with OUD and without OUD. **a**) The group with OUD (OUD+; orange) and without OUD (OUD-; blue) differed significantly on all measures shown (*t*-tests, all *p* < 9.36e-03). Shading of the orange and blue colors indicate the responses from “Not at all” to “Extremely”. Horizontal lines are centered around “Moderately”. The width of the bars indicates the proportion of individuals endorsing each response. **b**) Difference in mean response between OUD+ and OUD-. Responses were assigned a value such that “Not at all” was equal to 0 and “Extremely” was equal to 4. While the mean of most subjective responses was significantly higher in the OUD+ group, the mean of “Dizzy” and “Tired” were significantly lower.

The observation that initial positive subjective effects of opioids are strongly associated with OUD suggests the possibility that they could be used to predict risk for OUD. To investigate this idea, we used three classifiers (Logistic Regression, XGBoost, and Random Forest) to build prediction models for OUD (**Figure 2**), which we then evaluated using a ROC (**Figure 2a**), PR-AUC (**Figure 2b**), SHAP (**Figure 2c**), and OR (**Figure 2d**).

**Figure 2.**
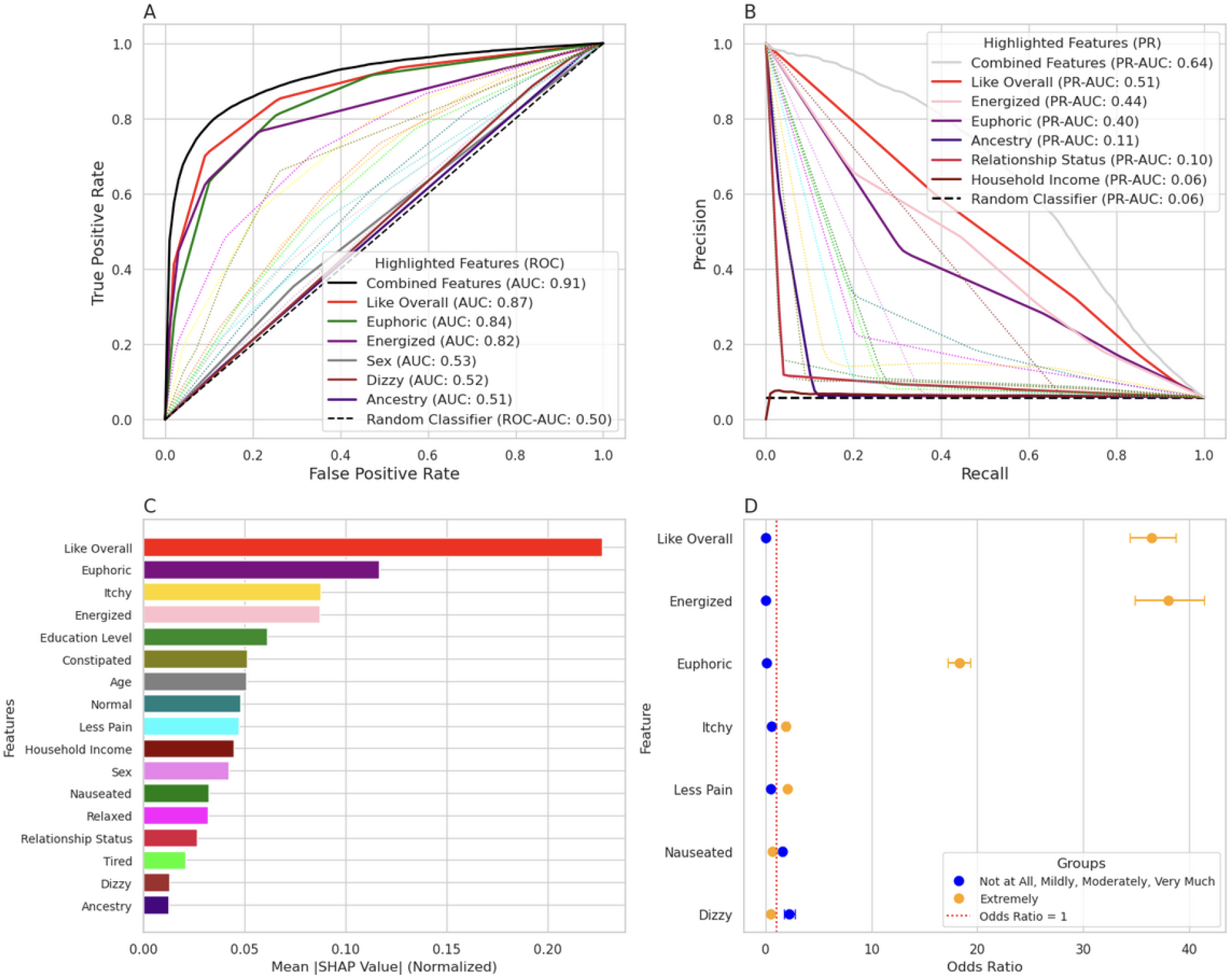
Initial subjective response to prescription opioids strongly predicted risk for opioid use disorder. **a)** Receiver operator characteristic (**ROC**) curves and their area under curves (**AUC**) comparing the predictive performance in terms of sensitivity and specificity for individual questions and their combinations. The dashed line represents a random classifier (ROC-AUC= 0.50). **b)** Precision-Recall (**PR**) curves for comparing the predictive performance of individual questions and their combinations. The dashed line represents a random classifier (PR-AUC = 0.06). **c)** Normalized mean SHapley Additive exPlanations (**SHAP**) values showing the relative importance of questions in the predictive model. **d)** Odds ratios for a subset of questions comparing “Extremely” (orange) to all other responses (blue). Error bars represent 95% confidence intervals. The red dashed line marks an odds ratio of 1. All results were statistically significant (*p* < 1.96e-12; **Table S4-S5**). Including covariates (age, sex, relationship status, ancestry, income, educational level) did not meaningfully alter the results (**Figure S6**).

**Figure 2a** illustrates the predictive performance of subjective effects and demographic features using ROC curves. The combined feature set, which incorporates all subjective effects and demographic variables, achieves the strongest performance (ROC-AUC = 0.91). Among individual features, “Like Overall” performs nearly as well (ROC-AUC = 0.87), followed by “Euphoric” (ROC-AUC = 0.84) and “Energized” (ROC-AUC = 0.82). In contrast, negative subjective effects like “Dizzy” (ROC-AUC = 0.52) were only slightly better than random performance. Similarly, demographic features such as “Sex” (ROC-AUC = 0.53) and “Ancestry” (ROC-AUC = 0.51) were also poor predictors of OUD status in the current sample.

PR curves (**Figure 2b**) further demonstrated the predictive value of positive subjective effects. The combined feature set achieved the highest PR-AUC (PR-AUC = 0.64). “Like Overall” maintained strong predictive performance (PR-AUC = 0.51), with “Energized” and “Euphoric” also performing well (PR-AUCs = 0.44 and 0.40, respectively). Negative subjective effects such as “Constipated” (PR-AUC = 0.13), provided little predictive value (**Table S4**). Demographic features like “Ancestry” (PR-AUC = 0.11) and “Sex” (PR-AUC = 0.23) also provided little predictive value (**Table S4**). Additional predictive performance metrics for all features and feature sets are provided in **Table S4**.

SHAP analysis (**Figure 2c**) also highlighted the relative importance of subjective effects in predicting OUD status. The positive “Like Overall” again emerged as the strongest predictor, with “Euphoric” and the negative “Itchy” also showing significant contributions.

Another way of quantifying the likelihood of reporting OUD is an OR (**Figure 2d**). The OR for responding “Extremely” for “Like Overall” and “Energized” were over 30, demonstrating an extraordinarily strong association between these questions and OUD. In contrast, negative subjective effects showed much smaller ORs (OR < 2).

Finally, we sought to use a decision tree classifier to identify the optimal set of questions to predict OUD status (**Figure 3**). We identified the two best questions for stratifying OUD risk. Consistent with the results shown in **Figure 2**, the response to “Like Overall” was the best predictor of OUD status. Although negative subjective effects generally had low predictive power (**Figure 2**), the optimal follow up question was “Itchy”. These findings highlight that the positive subjective effects question “Like Overall” is highly discriminative, with the inclusion of the negative subjective question “Itchy” further refining classification. These two questions separated the bulk of individuals (91,206; 64.3% of the cohort), who had just a 1.2% risk for OUD from others that had higher risk, including the highest risk group (3,188; 2.2% of the cohort) who had a 78.5% risk for OUD. Importantly, this decision tree reflects the fact that individuals who reported an unpleasant itchy feeling were at *higher* risk for OUD, showing that sensitivity to a negative effect confers greater risk, rather than being protective, as has been observed for sensitivity to the negative subjective effects of alcohol.^10,12^

**Figure 3.**
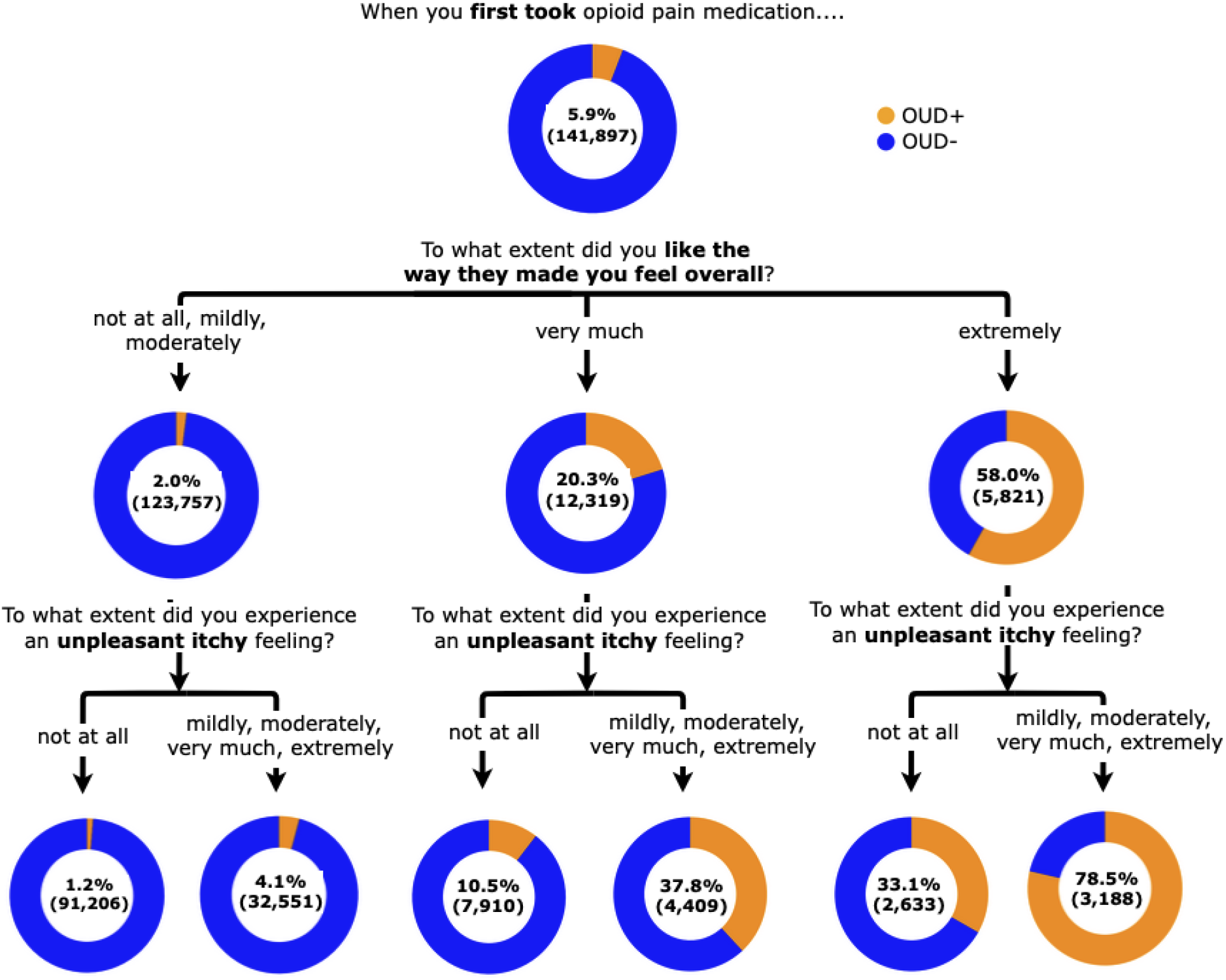
Optimal two-question decision tree for predicting risk for opioid use disorder. Orange represents individuals self-reporting OUD while blue represents those not reporting OUD. 5-fold validation yielded equivalent results.

## DISCUSSION

Using a large cohort of 141,897 participants, we show that *positive* initial responses to prescription opioids strongly predict the risk of OUD. Although opioids are prescribed for their analgesic properties, one novel finding of our study was that these effects were only slightly higher in individuals reporting OUD, suggesting that the analgesic effects, which could be construed as positive, are in fact dissociable from the positive effects that confer risk for OUD. This observation suggests that the differences in positive responses were not secondary to differences in overall sensitivity, metabolism, or differences in the dose or drug. Because of the size of our cohort, we were able to identify which individual questions best predicted OUD. The single question, “Like Overall”, was highly predictive of OUD. In fact, “Like Overall” was almost as predictive as using all available questions. Using the full dataset, we created an optimized 2-question decision tree that identifies a small subset who are at high risk for OUD. These two questions can easily be implemented in clinical settings where stratifying risk for OUD is urgently needed.

There has been some controversy in the addiction field about the relationship between initial responses to drugs and the risk for future use and misuse. One line of research with alcohol indicates that insensitivity to initial effects of alcohol is a risk factor for alcohol use disorder^10,12^, but more recent work suggests that subjective positive effects of alcohol are predictive of future problematic alcohol use.^8,13^ Similarly, positive subjective effects have also been identified as risk factors for misuse of stimulants^4,7,9^ and cannabis^5,6,11^. There is also prior evidence that initial positive subjective effects of opioids predict future problematic use. Double blind, laboratory-based experimental studies in healthy subjects demonstrated that pleasurable opioid effects predict whether the subject would choose the drug over placebo.^14,15^ Two retrospective studies examining initial responses to opioids showed that subjective effects differed in participants with and without OUD (*n* = 40) and in opioid misusing participants with and without OUD (*n* = 50).^28,29^ Once OUD developed, individuals with OUD were more likely to report positive subjective effects following acute morphine and heroin injections.^18,32,33^ However, it was not clear whether these individuals experienced more positive effects prior to developing OUD. Our study differs from these prior studies by using an extremely large population and in its use of that large dataset to identify the two questions that best predict risk for OUD.

To facilitate clinical implementation, we developed a two-question decision tree that stratifies individuals based on their risk for OUD. The most discriminatory question was “Like Overall” followed by sensitivity to itchiness. Surprisingly, individuals reporting more itchiness, which is a negative subjective effect, were at higher risk for OUD, contrasting with the alcohol literature in which sensitivity to negative effects is protective against AUD. While “Itchy” alone was a poor predictor of OUD (**Figure 2d**), when used in combination with “Like Overall” it further stratified risk for OUD (**Figure 3**). Prior literature has highlighted the importance of itchiness as a measure of greater sensitivity to opioids.^34–36^ Although other tools have been developed to estimate the risk of OUD, such as the Screener and Opioid Assessment for Patients with Pain (**SOAPP** and the **SOAPP-R**)^37,38^, and the ORT^20^ they are longer, have not been validated in large, non-ascertained populations and are not widely used. We performed simulations (**Figure S7**) that showed that our models could not have been optimized with a sample size lower than 40,000.

There are several major theories of the etiology of substance use disorders. One theory involves allosteric homeostasis, related to repeated cycles of intoxication and withdrawal^39^, whereas others relate more to the positive or hedonic effects of drugs.^40,41^ Our results support the idea that initial hedonic responses to opioids dramatically increase risk for OUD. Future research may suggest ways to integrate these apparently opposing theoretical frameworks. For example, it is possible that a stronger hedonic response in at-risk individuals triggers stronger opposing responses that lead to intensifying cycles of intoxication and withdrawal.

This study is not without limitations. First, we used 23andMe research participants who are likely to have numerous unique characteristics. Notably, they were disproportionately female, highly educated (88.61% with some college), higher income and reporting unusually high levels of multiple psychiatric diseases (e.g., 46.2% self-reported anxiety, 46.9% self-reported depression; **Table S2**). It remains to be seen to what extent our results can be generalized to other populations. Second, our study relies on retrospective self-reported data, which may be susceptible to recall bias. Individuals with OUD may have inaccurate recollection of their earliest experience with prescription opioids due to their long history of use. Future longitudinal studies could address this limitation. Third, we did not capture factors like the specific opioid, dose, or route of administration associated with the initial experience with opioids, leaving open the possibility that the difference in the positive subjective responses between the groups may reflect these factors. However, the fact that both groups reported similar levels of analgesia argues against this interpretation. Fourth, we propose that questions about subjective effects of opioids could be used to predict the risk for developing OUD. However, this approach will only be possible for patients who have previous experience with prescription opioids. In addition, this approach is less likely to be effective in opioid seeking patients, including those who have already developed OUD, since they may suspect that indicating that they like the subjective effects of opioids would make a physician reluctant to prescribe them. Finally, our study assumed that the first experience with opioids was intended to treat pain, how a participant might have responded to questions about pain if their first experience was purely recreational is unclear and could have confounded the responses to the analgesia question.

In summary, our findings show that initial sensitivity to the positive subjective effects of opioids are strongly related to OUD and we identified two questions that could be used to stratify risk for OUD. What to do once individuals have been stratified for OUD risk is a critical question for future studies. Individuals at higher risk may benefit from personalized counseling, reduced or different medications, or greater supervision.

## Data Availability

Due to participant privacy, data will not be available.

## ACKNOWLEDGEMENTS

This work was supported by National Institutes of Health grant numbers DP1DA054394 (JJM, SSR), R01DA061977 (AAP, SSR, JG), P50DA037844 (EOJ, AAP, SSR), T32GM139790 (JG) and R25MH081482 (LV-R).

We would like to thank the research participants and employees of 23andMe for making this work possible.

The following members of the 23andMe Research Team contributed to this study: Stella Aslibekyan, Adam Auton, Elizabeth Babalola, Robert K. Bell, Jessica Bielenberg, Ninad S. Chaudhary, Zayn Cochinwala, Sayantan Das, Emily DelloRusso, Payam Dibaeinia, Sarah L. Elson, Nicholas Eriksson, Chris Eijsbouts, Teresa Filshtein, Pierre Fontanillas, Davide Foletti, Will Freyman, Zach Fuller, Julie M. Granka, Chris German, Éadaoin Harney, Alejandro Hernandez, Barry Hicks, David A. Hinds, M. Reza Jabalameli, Ethan M. Jewett, Yunxuan Jiang, Sotiris Karagounis, Lucy Kaufmann, Matt Kmiecik, Katelyn Kukar, Alan Kwong, Keng-Han Lin, Yanyu Liang, Bianca A. Llamas, Aly Khan, Steven J. Micheletti, Matthew H. McIntyre, Meghan E. Moreno, Priyanka Nandakumar, Dominique T. Nguyen, Jared O’Connell, Steve Pitts, G. David Poznik, Alexandra Reynoso, Shubham Saini, Morgan Schumacher, Leah Selcer, Anjali J. Shastri, Jingchunzi Shi, Suyash Shringarpure, Keaton Stagaman, Teague Sterling, Qiaojuan Jane Su, Joyce Y. Tung, Susana A. Tat, Vinh Tran, Xin Wang, Wei Wang, Catherine H. Weldon, Amy L. Williams, Peter Wilton.

## REFERENCES

1. Butler, M. M. et al. Emergency Department Prescription Opioids as an Initial Exposure Preceding Addiction. Ann. Emerg. Med. 68, 202–208 (2016).

2. Edlund, M. J. et al. The role of opioid prescription in incident opioid abuse and dependence among individuals with chronic noncancer pain: the role of opioid prescription. Clin. J. Pain 30, 557–564 (2014).

3. Kendler, K. S., Lönn, S. L., Ektor-Andersen, J., Sundquist, J. & Sundquist, K. Risk factors for the development of opioid use disorder after first opioid prescription: a Swedish national study. Psychol. Med. 53, 6223–6231 (2022).

4. Davidson, E. S., Finch, J. F. & Schenk, S. Variability in subjective responses to cocaine: initial experiences of college students. Addict. Behav. 18, 445–453 (1993).

5. Davidson, E. S. & Schenk, S. Variability in subjective responses to marijuana: Initial experiences of college students. Addict. Behav. 19, 531–538 (1994).

6. Fergusson, D. M., Horwood, L. J., Lynskey, M. T. & Madden, P. A. F. Early Reactions to Cannabis Predict Later Dependence. Arch. Gen. Psychiatry 60, 1033–1039 (2003).

7. Haertzen, C. A., Kocher, T. R. & Miyasato, K. Reinforcements from the first drug experience can predict later drug habits and/or addiction: Results with coffee, cigarettes, alcohol, barbiturates, minor and major tranquilizers, stimulants, marijuana, hallucinogens, heroin, opiates and cocaine. Drug Alcohol Depend. 11, 147–165 (1983).

8. King, A. et al. Subjective Responses to Alcohol in the Development and Maintenance of Alcohol Use Disorder. Am. J. Psychiatry 178, 560–571 (2021).

9. Newton, T. F. et al. Bupropion Reduces Methamphetamine-Induced Subjective Effects and Cue-Induced Craving. Neuropsychopharmacology 31, 1537–1544 (2006).

10. Schuckit, M. A. Self-rating of alcohol intoxication by young men with and without family histories of alcoholism. J. Stud. Alcohol 41, 242–249 (1980).

11. Le Strat, Y. et al. First positive reactions to cannabis constitute a priority risk factor for cannabis dependence. Addiction 104, 1710–1717 (2009).

12. Schuckit, M. A. Low level of response to alcohol as a predictor of future alcoholism. Am. J. Psychiatry 151, 184–189 (1994).

13. King, A. C., Cao, D., deWit, H., O’Connor, S. J. & Hasin, D. S. The role of alcohol response phenotypes in the risk for alcohol use disorder. BJPsych Open 5, e38 (2019).

14. Zacny, J. P. Characterizing the subjective, psychomotor, and physiological effects of a hydrocodone combination product (Hycodan) in non-drug-abusing volunteers. Psychopharmacology (Berl.) 165, 146–156 (2003).

15. Zacny, J. P. & Lichtor, S. A. Within-subject comparison of the psychopharmacological profiles of oral oxycodone and oral morphine in non-drug-abusing volunteers. Psychopharmacology (Berl.) 196, 105–116 (2008).

16. Bieber, C. M. et al. Retrospective accounts of initial subjective effects of opioids in patients treated for pain who do or do not develop opioid addiction: a pilot case-control study. Exp. Clin. Psychopharmacol. 16, 429–434 (2008).

17. Agrawal, A. et al. Retrospectively assessed subjective effects of initial opioid use differ between opioid misusers with opioid use disorder (OUD) and those who never progressed to OUD: Data from a pilot and a replication sample. J. Neurosci. Res. 100, 353–361 (2022).

18. McAuliffe, W. E. A Second Look at First Effects: The Subjective Effects of Opiates on Nonaddicts. J. Drug Issues 5, 369–397 (1975).

19. Courchesne-Krak, N. S. et al. Prescription Opioid Medication Survey: A Tool to Collect Deep Phenotypic Data on the Multifactorial Pathways to Opioid Use Disorder in Clinical and Population-Based Cohorts. Complex Psychiatry 11, 72–93 (2025).

20. Webster, L. R. & Webster, R. M. Predicting Aberrant Behaviors in Opioid-Treated Patients: Preliminary Validation of the Opioid Risk Tool. Pain Med. 6, 432–442 (2005).

21. Butler, S. F. et al. Development and validation of the Current Opioid Misuse Measure. Pain 130, 144–156 (2007).

22. Coyne, K. S. et al. Construct validity and reproducibility of the Prescription Opioid Misuse And Abuse Questionnaire (POMAQ). Curr. Med. Res. Opin. 37, 493–503 (2021).

23. Knisely, J. S., Wunsch, M. J., Cropsey, K. L. & Campbell, E. D. Prescription Opioid Misuse Index: a brief questionnaire to assess misuse. J. Subst. Abuse Treat. 35, 380–386 (2008).

24. McLellan, A. T., Luborsky, L., Woody, G. E. & O’Brien, C. P. An improved diagnostic evaluation instrument for substance abuse patients. The Addiction Severity Index. J. Nerv. Ment. Dis. 168, 26–33 (1980).

25. Skinner, H. A. The drug abuse screening test. Addict. Behav. 7, 363–371 (1982).

26. Workman, T. E. et al. A Comparison of Veterans with Problematic Opioid Use Identified through Natural Language Processing of Clinical Notes versus Using Diagnostic Codes. Healthc. Basel Switz. 12, 799 (2024).

27. Palumbo, S. A. et al. Assessment of Probable Opioid Use Disorder Using Electronic Health Record Documentation. JAMA Netw. Open 3, e2015909 (2020).

28. Durand, E. Y. et al. A scalable pipeline for local ancestry inference using tens of thousands of reference haplotypes. Preprint at 10.1101/2021.01.19.427308 (2021).

29. Chen, T. & Guestrin, C. XGBoost: A Scalable Tree Boosting System. in Proceedings of the 22nd ACM SIGKDD International Conference on Knowledge Discovery and Data Mining 785–794 (Association for Computing Machinery, New York, NY, USA, 2016). doi:10.1145/2939672.2939785.

30. Pedregosa, F. et al. Scikit-learn: Machine Learning in Python. Mach. Learn. PYTHON (2011).

31. Lundberg, S. M. et al. From local explanations to global understanding with explainable AI for trees. Nat. Mach. Intell. 2, 56–67 (2020).

32. Lasagna, L., Felsinger, J.M.von & Beecher, H. K. Drug-induced mood changes in man: 1. observations on healthy subjects, chronically ill patients, and ‘postaddicts’. J. Am. Med. Assoc. 157, 1006–1020 (1955).

33. Von Felsinger, J. M., Lasagna, L. & Beecher, H. K. Drug-induced mood changes in man. II. Personality and reactions to drugs. J. Am. Med. Assoc. 157, 1113–1119 (1955).

34. Angst, M. S. et al. Aversive and Reinforcing Opioid Effects: A Pharmacogenomic Twin Study. Anesthesiology 117, 22–37 (2012).

35. Lipman, Z. M. & Yosipovitch, G. Substance use disorders and chronic itch. J. Am. Acad. Dermatol. 84, 148–155 (2021).

36. Reich, A. & Szepietowski, J. C. Opioid-induced pruritus: an update: Opioid-induced pruritus. Clin. Exp. Dermatol. 35, 2–6 (2010).

37. Butler, S. F., Budman, S. H., Fernandez, K. & Jamison, R. N. Validation of a screener and opioid assessment measure for patients with chronic pain. PAIN 112, 65 (2004).

38. Butler, S. F., Budman, S. H., Fernandez, K. C., Fanciullo, G. J. & Jamison, R. N. Cross-Validation of a Screener to Predict Opioid Misuse in Chronic Pain Patients (SOAPP-R). J. Addict. Med. 3, 66–73 (2009).

39. Koob, G. F. & Le Moal, M. Drug Addiction, Dysregulation of Reward, and Allostasis. Neuropsychopharmacology 24, 97–129 (2001).

40. Robinson, T. E. & Berridge, K. C. The neural basis of drug craving: an incentive-sensitization theory of addiction. Brain Res. Brain Res. Rev. 18, 247–291 (1993).

41. Wise, R. A. & Bozarth, M. A. A psychomotor stimulant theory of addiction. Psychol. Rev. 94, 469–492 (1987).

