## Supplemental Material for "How it begins: Initial response to opioids strongly predicts self-reported opioid use disorder"

**Exclusion Criteria**

Beyond the exclusion criteria described in the main Methods we also applied further exclusion criteria. Participants who responded with a preference for $20 to the question "*Would you rather have: $60 Today or $20 Today?*” were excluded from the analysis due to our belief that they were answering the survey carelessly.^1^ We also filtered out those who claimed to have first used opioids at an age older than their current age. Further, we removed anyone whose age at first use was below 12 years old due to the belief that a participant who first used opioids may not remember their effects. We also removed anyone currently below the age of 18 or above the age of 90 due to similar beliefs. The number of people excluded, and percent of total are reported in **Figure S2**. The final sample consisted of individuals who provided complete responses to all demographic, subjective effects and problematic opioid use questions (*n* = 141,897).

**Statistical Analyses**

We ran a sensitivity analysis among the four groups defined by combinations of OUD diagnosis (OUD+ vs. OUD−) and problematic opioid use score (high vs. low), comparing subjective effect responses using ANOVA followed by Tukey post-hoc tests. This analysis was used to validate the empirical cut-offs for excluding participants whose OUD status was discordant with their problematic opioid use score (**Figure S4**).

We sought to use the responses to the subjective effects questions to predict OUD status using three classifiers: Logistic Regression, XGBoost, and Random Forest with default hyperparameter settings. We implemented Logistic Regression and Random Forest with implemented using the *sklearn*^2^ package and XGBoost with the *xgboost package*^3^.

We evaluated the models' performance, using a 5-fold cross-validation and averaging metrics across the three classifiers. We then calculated true and false positive rates across thresholds and plotted them on a receiver operating characteristic (**ROC**) curve. We also calculated the area under the curve (**AUC**), which summarizes the model’s ability to distinguish between the two classes. Further, we calculated precision (also known as positive predictive value which measures the proportions of positive predictions to the self-reported OUD status) and recall (same as true positive rate) across thresholds and visualized the results using a precision-recall (**PR**) curve. The PR curve emphasizes the trade-off between precision and recall, which is particularly informative given the imbalanced nature of our dataset. The area under the PR curve (**PR-AUC**) provided a summary measure of the model's ability to identify OUD status while minimizing false positives. We also calculated the F1 score, the harmonic mean of precision and recall. These metrics were calculated on various feature sets including ones that were limited to individual subjective effects, individual demographics, and the combination of both feature sets. SHapley Additive exPlanations (**SHAP**)^4^ values were then calculated for each of the classifiers then normalized and averaged together. Odds ratios (**OR**) were then calculated for a subset of the features, comparing those answering the extreme value of “Extreme” to those who answered anything else. We also ran our models on features with demographics as covariates, which yielded similar results to the one reported in the manuscript (**Figure S6**).

We trained a decision tree classifier using *sklearn*^2^ to identify the minimal set of questions relevant to OUD. The decision tree was fit using the same combined feature set described above, with a maximum depth of three and a minimum impurity decrease of 0.001. We also created 5 trees with subsets of the data using the same 5-fold cross validation method as was used in the classifiers to test stability, each yielded equivalent results to the one reported in the manuscript.

To determine the minimum sample size required for stable prediction of problematic opioid use risk risk, we conducted a subsampling analysis. Stratified random subsets ranging from 0 to 60,000 participants were used to train models, each evaluated using 5-fold cross-validation across 5 random replicates. We computed average performance metrics including ROC-AUC, PR-AUC, precision, recall, and F1-score at each sample size (**Figure S7**). Performance steadily improved with increasing sample size and began to stabilize beyond approximately 40,000 participants.

All analyses were completed in Python 3.11.

**REFERENCES**

1.	Courchesne-Krak NS, Chandrasekaran A, Gonzalez J, et al. Prescription Opioid Medication Survey: A tool to collect deep phenotypic data on the multifactorial pathways to opioid use disorder in clinical and population-based cohorts. Complex Psychiatry; 2025. 11 (1): 72-93.

2.	Pedregosa F, Pedregosa F, Varoquaux G, et al. Scikit-learn: Machine Learning in Python. Mach Learn PYTHON 2011

3.	Chen T, Guestrin C. XGBoost: A Scalable Tree Boosting System. In: Proceedings of the 22nd ACM SIGKDD International Conference on Knowledge Discovery and Data Mining. New York, NY, USA: Association for Computing Machinery; 2016. p. 785-94

4.	Lundberg SM, Lee S-I. A Unified Approach to Interpreting Model Predictions [Internet]. In: Advances in Neural Information Processing Systems. Curran Associates, Inc.; 2017

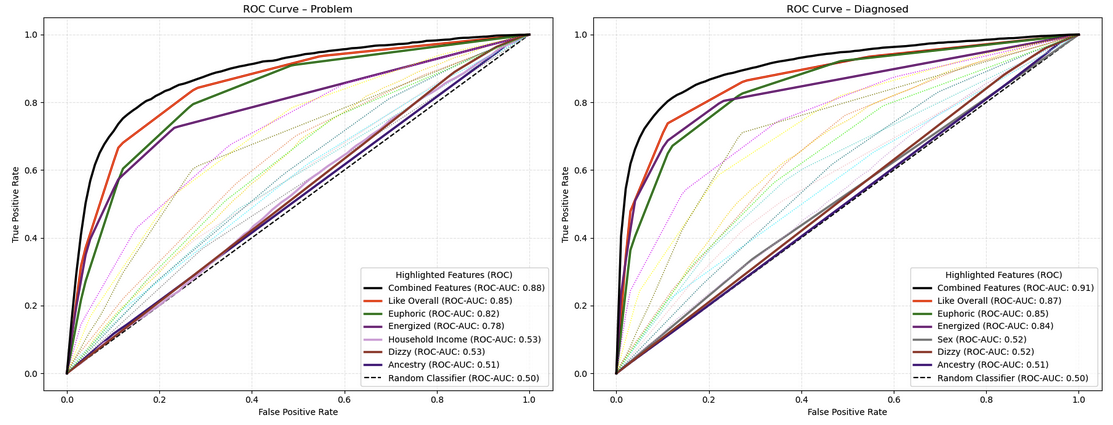

**Figure S1.** **Predictive performance across groups**. Receiver operator characteristic (**ROC**) curves and their area under curves (**AUC**) comparing the predictive performance in terms of sensitivity and specificity for individual questions and their combinations for both self-reported OUD problem and self-reported diagnosed opioid use disorder. The dashed line represents a random classifier (ROC-AUC= 0.50).

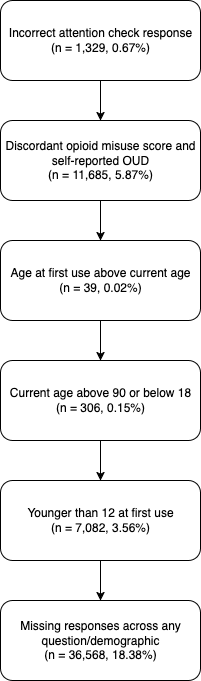

**Figure S2. Exclusion criteria and number of participants excluded at each step.**

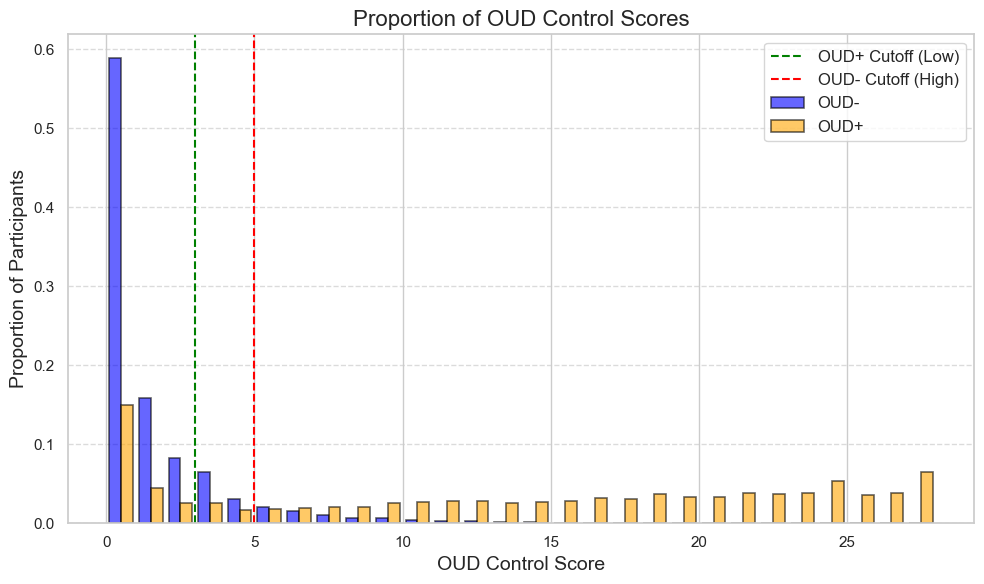

**Figure S3.** **Histogram of opioid misuse total scores in individuals self-reporting OUD (OUD+) and those without (OUD-)**. To minimize misclassification errors, participants were excluded if their total problematic opioid use score was discordant with their response to the self-reported lifetime OUD question. Specifically, we excluded participants who indicated they had OUD but whose total problematic opioid use score was below 3 (*n* = 2,442, 1.2%). We also excluded participants who responded that they did not have OUD but whose total problematic opioid use score was above 5 (*n* = 9,243, 4.6%).

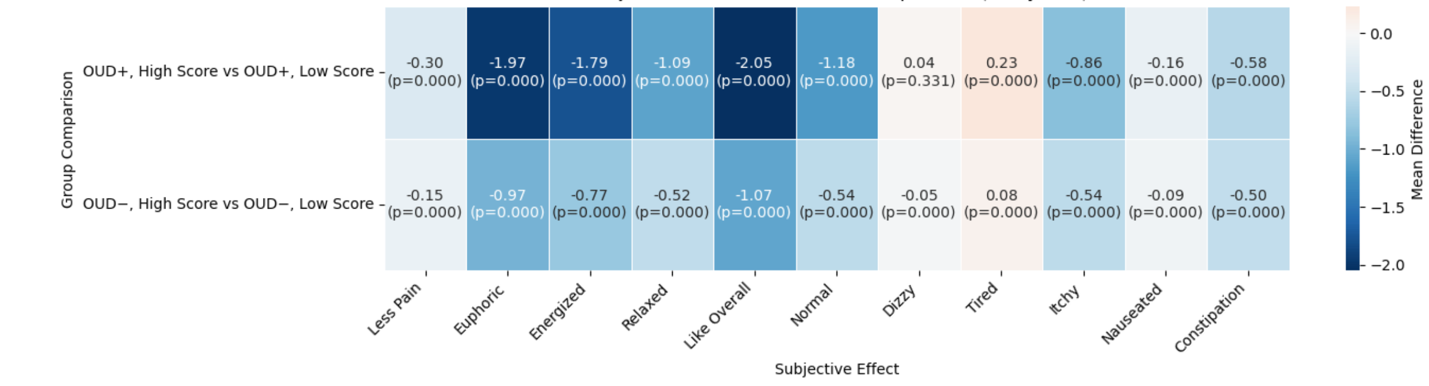

**Figure S4.** **Differences in subjective opioid responses between high and low problematic opioid use risk groups, stratified by OUD diagnosis**. Each cell shows the mean difference in reported subjective effect ratings between individuals with high vs. low total opioid misuse scores within OUD+ and OUD− groups. Positive values indicate greater endorsement among high-risk individuals. Group differences were assessed using ANOVA followed by Tukey post-hoc tests; all differences were statistically significant except for the “*Dizzy*” item in the OUD+ group.

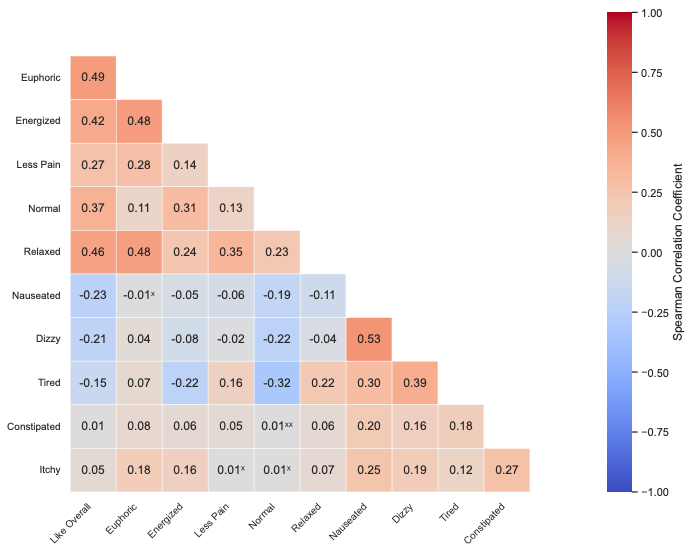

**Figure S5.** **Spearman correlation estimates (ρ) for all subjective effects.** Cells marked with **"xx"** indicate correlations that were not statistically significant (p > 0.05), while cells marked with **"x"** indicate correlations that did not meet the Bonferroni significance threshold (p > 9.1e-4).

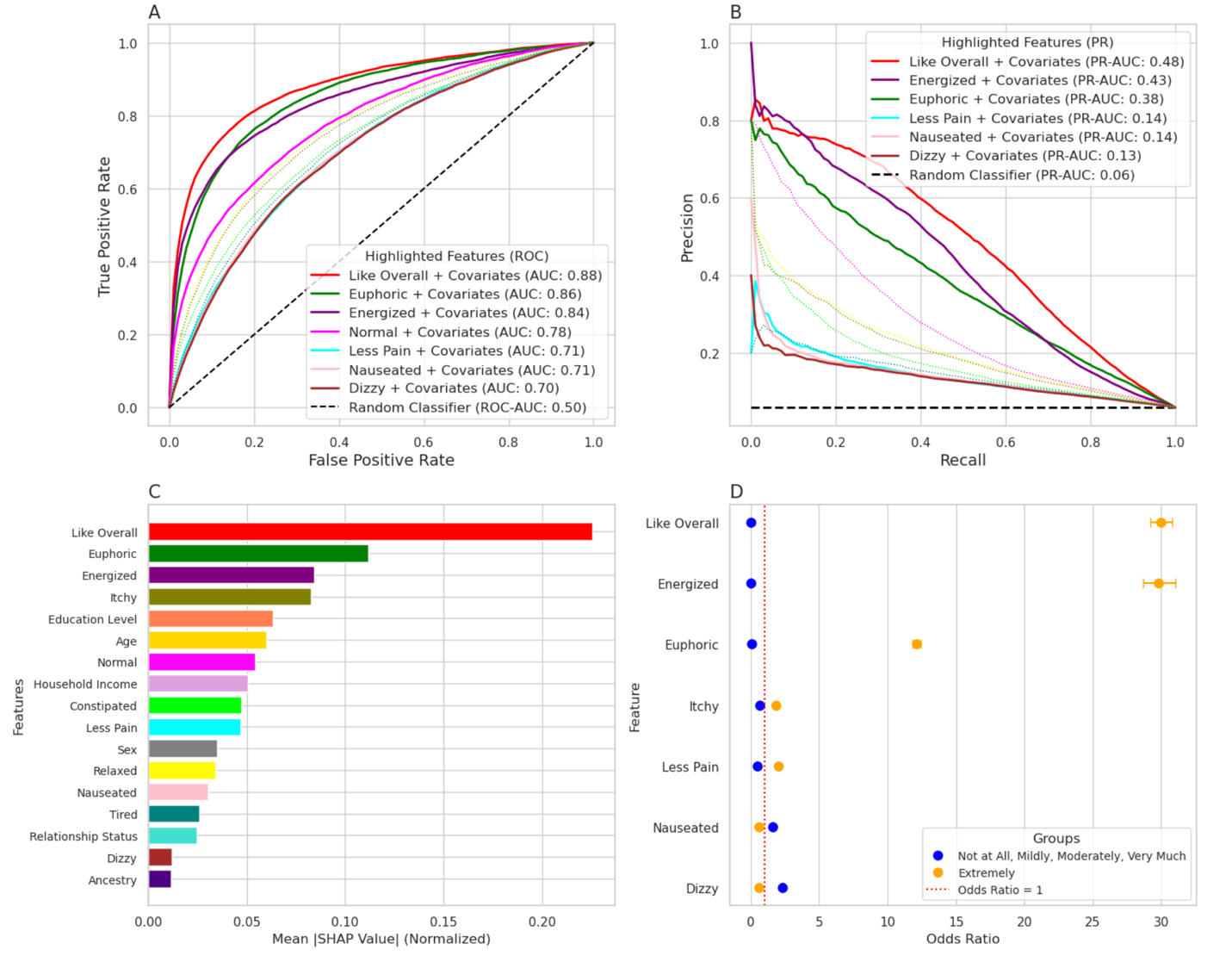

**Figure S6. Initial Subjective Response to Prescription Opioids Strongly Predicts Risk for Opioid Use Disorder Even with the Addition of Covariates (age, sex, relationship status, ancestry, income, educational level). a)** Receiver operator characteristic (**ROC**) curves and their area under curves (**AUC**) comparing the predictive performance in terms of sensitivity and specificity for individual questions and their combinations. The dashed line represents a random classifier (ROC-AUC= 0.50). **b)** Precision-Recall (**PR**) curves for comparing the predictive performance of individual questions and their combinations. The dashed line represents a random classifier (PR-AUC = 0.05). **c)** Normalized mean SHapley Additive exPlanations (**SHAP**) values showing the relative importance of questions in the predictive model. This remains the same compared to the original model as SHAP includes all covariates in the model regardless. **d)** Odds ratios for a subset of questions comparing “Extremely” (orange) to all other responses (blue). Error bars represent 95% confidence intervals. The red dashed line marks an odds ratio of 1.

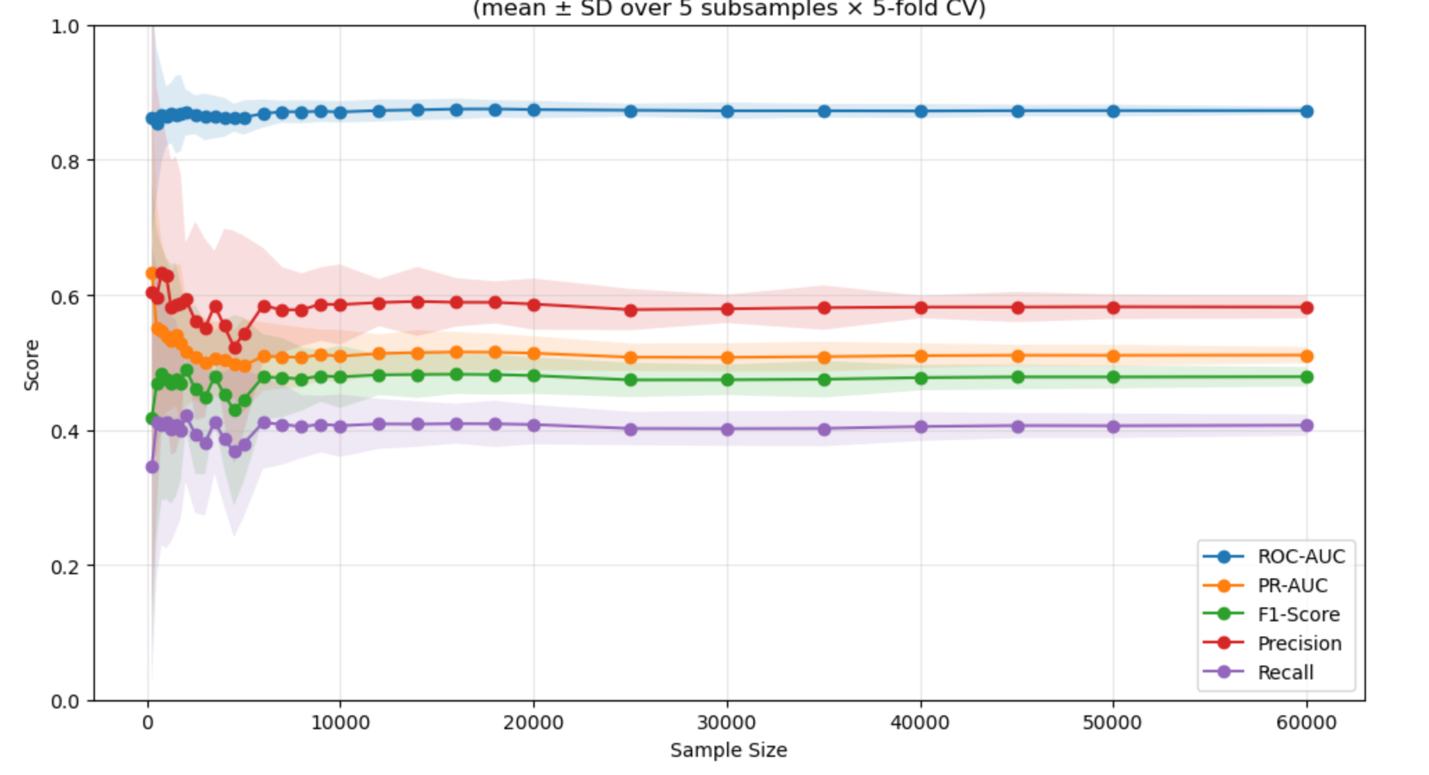

**Figure S7.** **Model performance across increasing sample sizes for predicting problematic opioid use risk.** Using 5-fold cross-validation averaged over 5 random subsamples per size, we observed that ROC-AUC remained consistently high, while PR-AUC, precision, recall, and F1-score steadily improved with increasing sample size and began to stabilize beyond approximately 40,000 participants.
